# Unsupervised clustering reveals a unique Treg profile in slow progressors to type 1 diabetes

**DOI:** 10.1101/2021.01.13.21249751

**Authors:** Joanne Boldison, Anna E Long, Rachel J Aitken, Isabel V Wilson, Claire Megson, Stephanie J Hanna, F. Susan Wong, Kathleen M Gillespie

## Abstract

**Objective:** To profile CD4^+^ regulatory T cells (Tregs) in a well-characterised cohort of slow progressors to type 1 diabetes, individuals positive for multiple islet autoantibodies who remain diabetes-free for at least 10 years.

**Research Design and Methods:** Peripheral blood samples were obtained from extreme slow progressor individuals (n=8), with up to 32 years follow-up, and age and gender-matched to healthy donors. One participant in this study was identified with a raised HbA1c at the time of assessment, and was individually evaluated in the data analysis. PBMCs were isolated, from donors, and to assess frequency, phenotype and function of Tregs, multi-parameter flow cytometry and T cell suppression assays were performed. Unsupervised clustering analysis, FlowSOM and CITRUS, was used to evaluate Treg phenotypes.

**Results:** Treg mediated suppression of CD4^+^ effector T cells, from slow progressors was significantly impaired, compared to healthy donors (P<0.05). Effector CD4 T cells, from slow progressors, were more responsive to Treg suppression, compared to healthy donors, demonstrated by increased suppression of CD25 expression on effector CD4 T cells (P<0.05). Unsupervised clustering on memory CD4 T cells, from slow progressors, showed an increased frequency of activated-memory CD4 Tregs associated with increased expression of GITR, compared to healthy donors (P<0.05). The participant with a raised HbA1c had a different Treg profile, compared to slow progressors and the matched controls.

**Conclusions:** CD4^+^ Tregs from slow progressor individuals have a unique Treg signature. This report highlights the need for further study of Treg heterogeneity in individuals at-risk of developing type 1 diabetes.

## Introduction

Progression rates from the first appearance of islet autoantibodies to development of clinical symptoms of type 1 diabetes are well described in childhood, with 70% of multiple islet autoantibody-positive children developing diabetes within 10 years of seroconversion and this increases to 84% for those followed for 15 years (1). In contrast, the mechanisms underlying adult-onset type 1 diabetes, which represents more than half of clinical type 1 diabetes is under-investigated. Some multiple islet autoantibody-positive individuals progress more slowly and develop adult-onset type 1 diabetes, however others will remain at risk for decades. We previously described a small but very well characterised group of extreme slow progressors (2) who remained diabetes-free for at least 10 years after the first multiple islet autoantibody sample was detected. Subsequently, we showed that islet autoantigen-specific CD8^+^ T cell responses were largely absent in slow progressors but were readily detectable in individuals with recent-onset and longstanding diabetes (3). This might suggest that either regulation of the autoimmune response is enhanced in these individuals compared with those who progress, or that these individuals have been wrongly classified as “at risk” and are effectively heathy controls. To further understand if slow progressors have enhanced autoimmune regulation, we directed our study at evaluating regulatory T cells (Tregs).

Earlier studies have shown that although Tregs appear to be normal in number, individuals with diabetes have some functional defects, which include a reduced capacity to respond to IL-2 (4). In addition, effector CD4^+^ T cells in those who develop diabetes may be more resistant to regulation, demonstrated by a reduction in suppression of effector T cells, by both naturally occurring T regulatory cells, as well as *in vitro* generated adaptive T regulatory cells (5) and diminished IL-2 responsiveness in antigen experienced CD4^+^ T cells (6). The aim of this study was to examine whether Treg function, in this well-characterised cohort of extreme slow progressors (including an individual on the verge of diagnosis after 32 years follow up), can be differentiated from age- and gender-matched healthy control donors.

## Research Design and Methods

### Participants

The Bart’s Oxford (BOX) study is a longitudinal, population-based study examining risk factors for type 1 diabetes in relatives of probands diagnosed under the age of 21 years (7) and recruitment has been ongoing since 1985. We previously described the characteristics of long term slow progressors who remained multiple islet autoantibody positive for more than 10 years but did not develop clinical symptoms of diabetes (2). This included data on 36 slow progressors from the BOX study. Subsequently, 10 slow progressors who continued to remain diabetes-free and were willing to provide large volume blood samples were included in an analysis of T and B cell function (3). In the current study, 8 Slow Progressors, with median age 43 (range 31-72 years), had been islet autoantibody antibody positive for between 18 and 32 years (see supplementary table 1). The participant aged 72 years (ID: SP 606) was identified with a raised HbA1c in February 2020, which was subsequently confirmed by his Clinical team and he was formally diagnosed with diabetes 8 months after the sample taken for this study. The HbA1c for his sample analysed in this study was 53 mmol/mol.

### Ethics

The BOX study was approved by the South Central–Oxford C National Research Ethics Committee. Study of progressors and control individuals was approved by the South East Wales Research Ethics Committee and conducted in accordance with the principles of Good Clinical Practice established by the International Council for Harmonization/WHO. All participants provided written informed consent prior to enrolment, as mandated by the Declaration of Helsinki.

### Autoantibodies

Autoantibodies were measured using well described radioimmunoassays (8). For ZnT8A, two separate assays were performed for the variants R325 and W325, and the higher value used to determine response. The results were expressed in units derived from in-house standard curves for IAA and ZnT8A, measured in 5 μl and 2 μl of serum, respectively. For GADA and IA-2A, results were derived from a standard curve developed for the NIDDK-sponsored Islet Autoantibody Harmonization Program and were expressed in digestive and kidney (DK) units/ml (9). These assays were submitted to the Islet Autoantibody Standardisation Programme 2020 where they achieved sensitivity and specific of GADA – 64% & 97.8%, IA-2A – 72% & 100%, IAA – 60% & 97.8%, and ZnT8 – 68% & 100% (when combining results of variants as described), respectively.

### Peripheral Blood Samples

PBMCs were isolated from heparinised samples of whole blood via density gradient centrifugation using Lymphoprep (STEMCELL Technologies, Cambridge, UK). 1×10^6^ fresh PBMCs were taken for CD4 regulatory T cell flow cytometric analysis. Remaining PBMCs were used for CD4^+^ T cell suppression assays.

### Flow cytometry

Fresh PBMCs were incubated with TruStain (anti-human CD16/32 [Biolegend]) for 10min at 4°C, followed by fluorochrome-conjugated mAbs against cell surface markers for 30min at 4°C. Multi-parameter flow cytometry was carried out using mAbs: CD19 BV605 (HIB19), CD8 BV605 (HIT8a), CD4 AF700 (A161A1), CD25 PeCy7 (BC96), CD127 PerCPCy5.5 (A019D5), CD45RO BV421 (UCHL1), CD45RA APC-Cy7 (HI100), CD39 PE/DAZZLE 954 (A1), Lag-3 BV786 (11C3C65), CD49b FITC (P1E6-C5), GITR BV711 (108-17), HLA-DR BV650 (L243), FoxP3 AF647 (206D), CTLA4 PE (L3D10). Dead cells were excluded from analysis by Live/Dead exclusion dye (Invitrogen, MA, USA). For the transcription factor FoxP3 and CTLA4 expression, for which intracellular staining was performed, the cells were fixed/permeabilized using eBioscience nuclear transcription kit. Cells were acquired on LSRFortessa (FACS Diva software) and analysis was performed using FlowJo software (Treestar) and unsupervised clustering methods.

### Unsupervised clustering using FlowSOM and CITRUS

Initial data processing was performed on FlowJo. 10,000 events of live CD19^-^CD8^-^ CD4^+^CD45RA^-^ T cells, from each donor, were down-sampled and imported into R package and the FlowSOM algorithm was used define 10 metaclusters for phenotyping analysis. Clusters were visualised using both a minimum spanning tree (MST), and tSNE plots (t-distribution stochastic neighbour embedding). Heatmaps were generated using R package ggplot2. CITRUS (cluster identification, characterisation and regression) was performed using Cytobank. Samples were loaded in Cytobank and traditional gating performed on live CD8^-^CD19^-^CD4^+^CD45RA^-^ cells. Clustering was performed using equal event sampling (8000), and a minimum cluster size set to 1%. A false discovery rate (FDR) was 1%. For both FlowSOM and CITRUS the following markers were included: CD25, CD127, CD39, FoxP3, CTLA4, HLA-DR, GITR, Lag-3, CD49b.

### CD4^+^ T cell suppression assay

CD4^+^ T cells were isolated by magnetic activated cell sorting (MACS) using the CD4^+^CD25^+^ T cell isolation kit (Miltenyi), according to the manufacturer’s instructions. Briefly, CD4 T cells, from PBMCs, were negatively isolated. Total CD4 T cells were then separated into CD4^+^CD25^-^ and CD4^+^CD25^+^ fractions by positive selection using anti-CD25 microbeads. CD4^+^CD25^-^ (4×10^5^/well) responder cells were cultured with CD4^+^CD25^+^ at various ratios indicated. All CD4^+^CD25^-^ responder cells were labelled with CFDA-SE (CFSE) (Invitrogen) prior to culture set up. All co-cultures were stimulated with Dynabeads™ anti-CD3/28 beads (Invitrogen) (0.35μl beads/4×10^5^ cells, 1 bead: 28responders) and cultured for 3 days before analysis by flow cytometry with CD4 AF700, CD25 PeCy7. Dead cells were excluded from analysis by Live/Dead exclusion dye (Invitrogen, MA, USA). All cells were cultured in RMPI complete media containing 2mM L-glutamine, 100 U/mL Penicillin and 10% heat-inactivated AB serum (Sigma-Aldrich). Percent suppression was calculated by (% CFSE (or CD25) expression in Tresponder and Treg co-cultures)/(% CFSE (or CD25) in Tresponder alone cultures) × 100. Supernatants were taken from co-cultures at the 3-day endpoint, to analyse IFNγ and IL-17A cytokines, which were measured by a Meso Scale Discovery (MSD) system. MSD was performed according to the manufacturer’s instructions and evaluated using an MSD Sector Imager 6000.

### Statistics

Statistical analyses were performed using GraphPad Prism 8 (GraphPad Software, San Diego, CA). Significance was determined by Two-way ANOVA, followed by a Bonferroni post-test for more than two variables, and Wilcoxon matched-pairs signed rank test was performed for only two variables. Data were considered significant at p<0.05.

## Results

### CD4 Tregs show impaired suppressive capacity, but CD4 effector cells are more responsive to suppression in slow progressors

To test the functionality of CD4^+^CD25^+^ Tregs in our slow progressor (SP) cohort we utilised a well-established *in vitro* T cell suppression assay (10) (Figure 1). Slow progressors were age- and gender-matched to healthy donors (HD), and Treg-mediated suppression of CD4^+^ responder T cells (CD4^+^CD25^-^ T cells) was assessed by proliferation (CFSE) and activation (CD25). CD4^+^ responder T cells were labelled with CFSE, co-cultured with varying ratios of CD4^+^ Tregs and stimulated with anti-CD3/28 activation beads. For representative flow cytometric plots, see Supplementary Figure 1.

**Figure 1.**
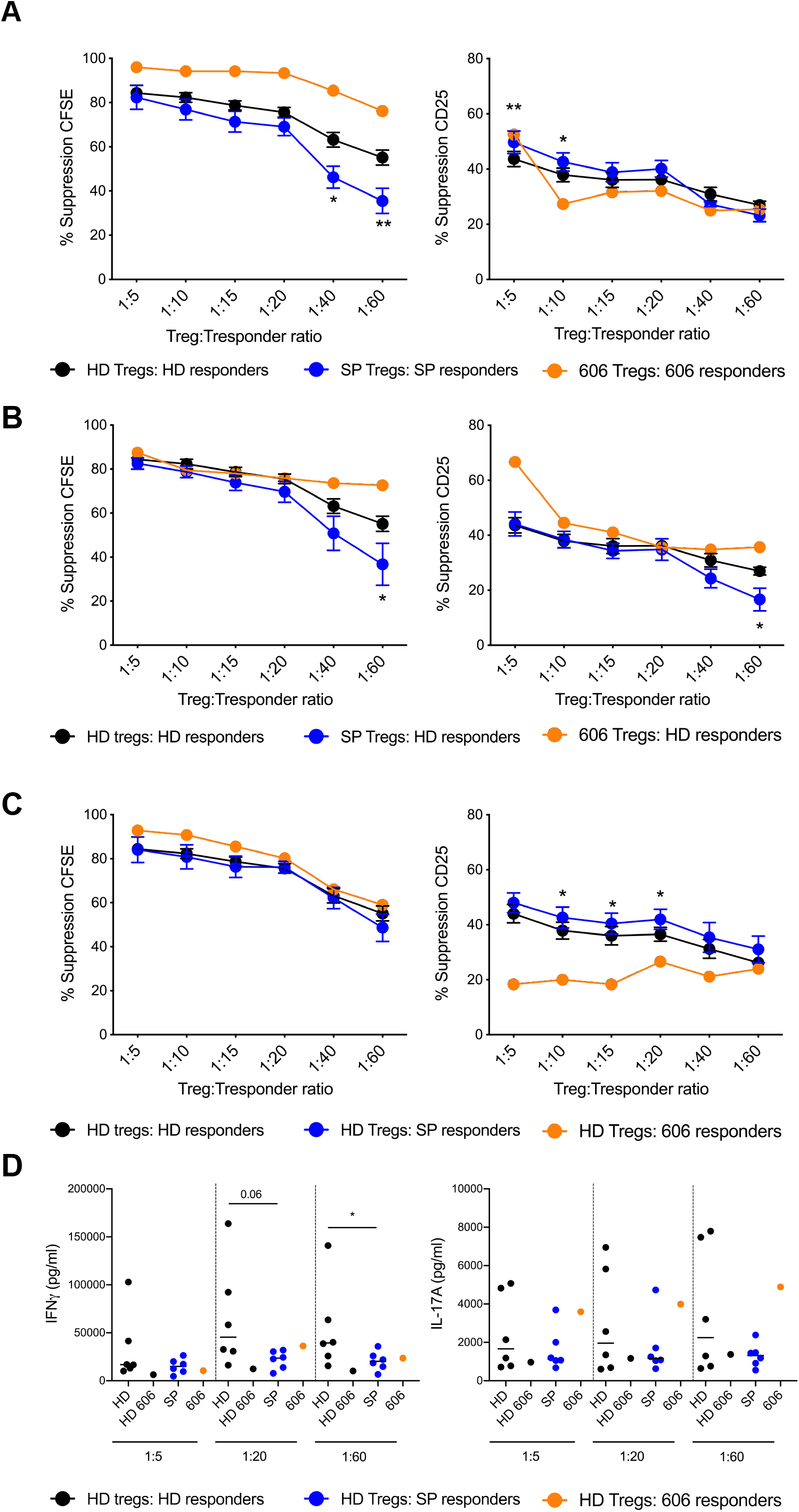
CD4^+^ Tregs from Slow Progressors are impaired, but effector CD4^+^ T cells are more responsive to suppression. Slow progressors (SP, blue line) were age- and gender-matched to healthy donors (HD, black line). The orange line represents slow progressor 606 with a raised HbA1c. CD4 T cells were sorted using a CD4^+^CD25^+^ Treg sorting kit. CD4^+^CD25^-^ (responders) were CFSE-labelled and Tregs were titrated at the observed ratios. Cells were activated with anti-CD3/28 beads and cultured for 3 days before flow cytometry (CD25 counterstain) and cytokine analysis. Percentage suppression was calculated using the positive control (activated responder cells with no Tregs). (A, B, C) Percentage Treg suppression calculated from CFSE proliferation (left) and CD25 (right) when co-cultures are (A) Tregs cultured with their CD4^+^ T cell responder counterparts (autologous) (B) Tregs from HD, SP or SP 606 are cultured with HD responders (C) HD Tregs are cultured with either HD, SP or SP 606 responders (D) IFNγ expression (left) and IL-17A (right) in co-cultures when HD Tregs are cultured with responders from matched HD (HD 606 is plotted separately), SP or SP donor 606. **<0.01; *<0.05, two-way ANOVA.

Firstly, we investigated autologous co-cultures to assess overall differences in Treg functionality (Figure 1A). Treg mediated suppression of CD4^+^ responders, as measured by CFSE proliferation, from slow progressors was significantly decreased compared to healthy donors (p<0.05) (Figure 1A, left). However, increased suppression of CD4^+^ T cell activation was observed at the higher Treg: Tresponder ratios (Figure 1A, right). Interestingly, Tregs from slow progressor 606 (53 mmol/mol HbA1c at the time of assessment) (orange line) suppressed CD4^+^ responders considerably more than healthy donors. It should be noted that no overall difference was observed in the levels of activation via anti-CD3/CD28 beads (measured both by CFSE proliferation and CD25 activation), in the absence of Treg, in slow progressors compared to healthy donors (data not shown).

Secondly, considering that the diminished suppression level in slow progressors could be attributed to either reduced suppressive ability of CD4^+^ Tregs or reduced responsiveness of CD4^+^ T responders to suppression in the co-culture, we performed allogeneic crossovers (Figure 1B, C). We assessed the suppressive ability of Tregs in co-cultures, with HD responders and Tregs from slow progressors. This revealed that slow progressor Tregs demonstrated significantly reduced suppression of both proliferation and activation of the CD4^+^ T cell responders, at a ratio of 1:60 Treg:Tresponder ratio (p<0.05), compared to HD Tregs (Figure 1B). This indicates that slow progressor Tregs are impaired in their ability to suppress CD4^+^ T cells, compared to matched healthy control donors.

Thirdly, we assessed CD4^+^ T cell responders and their resistance to suppression. We found that healthy donor Tregs co-cultured with slow progressor responders suppressed proliferation similarly to healthy donor responders (Figure 1C, left). However, suppression of CD25 on slow progressor CD4^+^ T cell responders, was significantly increased, compared to HD. Strikingly, CD4^+^ T cell responders from slow progressor 606 (orange line) were more resistant to suppression, as shown by CD25 measurement (Figure 1C, right). To confirm our observations in Figure 1C, we analysed pro-inflammatory cytokines IFNγ (Figure 1D, left) and IL-17A (Figure 1D, right). IFNγ expression was significantly reduced in co-cultures of healthy donor Tregs with slow progressor responders, compared to HD responders at a 1:60 Treg:Tresponder ratio. IL-17A production in the co-cultures was variable; however there was strikingly elevated IFNγ and IL-17A in the co-cultures with donor 606 responders (orange dots), compared to the matched HD. No significant differences in pro-inflammatory cytokines were observed when HD responders and SP Tregs were co-cultured (Supplementary Figure 2). In line with our autologous co-culture data in Figure 1A, we observed a significant decrease of IFNγ at the 1:20 Treg:Tresponder ratio, but no significant differences in IL-17A production (Supplementary Figure 3). Taken together, CD4^+^ Tregs from slow progressors show impaired suppressive capacity towards effector CD4^+^ T cells; however, CD4^+^ effector T cells from slow progressors are more responsive to Treg mediated suppression of T cell activation.

### Slow Progressors have significantly increased proportions of CD4^+^ effector memory regulatory CD4^+^ T cells (Tregs) compared to healthy donors

Next, we investigated the frequency of Tregs present in the peripheral blood compartment of slow progressors, compared to age- and gender-matched healthy donors. In our analysis we included both CD45RA and CD45RO surface markers to identify resting Tregs and activated effector Tregs, respectively. Overall, we found no differences in the percentages of CD4^+^CD45RA^+^ and CD4^+^CD45RO^+^ compartments in slow progressors compared to healthy donors (Supplementary Figure 4A). Furthermore, there was no statistically significant difference in CD4^+^CD45RA^+^CD25^+^CD127^lo^ resting Tregs (Supplementary Figure 4B). However, 6 out of 7 slow progressors had increased CD4^+^CD45RO^+^CD25^+^CD127^lo^ effector memory Tregs and this was significant (p<0.05) when compared to the healthy donor counterparts, including slow progressor 606 (orange square) (Supplementary Figure 4B). Furthermore, consistent with previous reports (11), CD25^+^ Tregs increased with age using our healthy donor cohort we observed a positive correlation (data not shown).

As Tregs are heterogeneous and there are a number of different subtypes (12), we chose to use unsupervised gating using FlowSOM (13), which is an automated algorithm based visualisation approach. Surface markers used for analysis included CD25, CD127, FOXP3, HLA-DR, CD39, CTLA4, GITR, CD49b, LAG3 and FlowSOM clustering was performed on down-sampled CD45RA^-^ cells that had been manually gated. FlowSOM identified 10 distinct clusters (Figure 2), which were identified as either as a memory Treg cell type or a memory T cell, based on expression of key markers (Figure 2B). CD4^+^ memory Treg cells were separated into 5 clusters, each characterised by individual phenotypes, with the most dominant being Memory Treg_3 and HLA-DR^+^GITR^+^ clusters (Figure 2C). Memory Treg clusters 1-4 had an intermediate to high expression of CD25 and FOXP3; however, CD39, HLA-DR and GITR were expressed heterogeneously. Analysis on each cluster from our slow progressor cohort revealed a significant increase in Memory Treg_3, compared to the age- and gender-matched healthy donors (p<0.01). Memory T cell_4 and HLA-DR^+^GITR^+^ clusters also increased in slow progressor individuals, but these increases did not reach statistical significance. Consistent with manual gating, slow progressor 606 (Figure 2C, orange dots) had an increase in memory Tregs compared to the matched healthy donor. Interestingly, we found a modest decrease in memory T cell_1 cluster in the slow progressors, compared to healthy donors (Figure 2C, red), supporting our previous observations (3). To assist in visualisation of the FlowSOM clusters, t-SNE plots were generated, overlaying each cluster identified for healthy donors and slow progressor cohorts (Figure 2D). t-SNE plots revealed memory Treg subsets clustered into distinct regions; however, some smaller subsets did not form discrete populations, indicating that these smaller subsets are part of a larger population.

**Figure 2.**
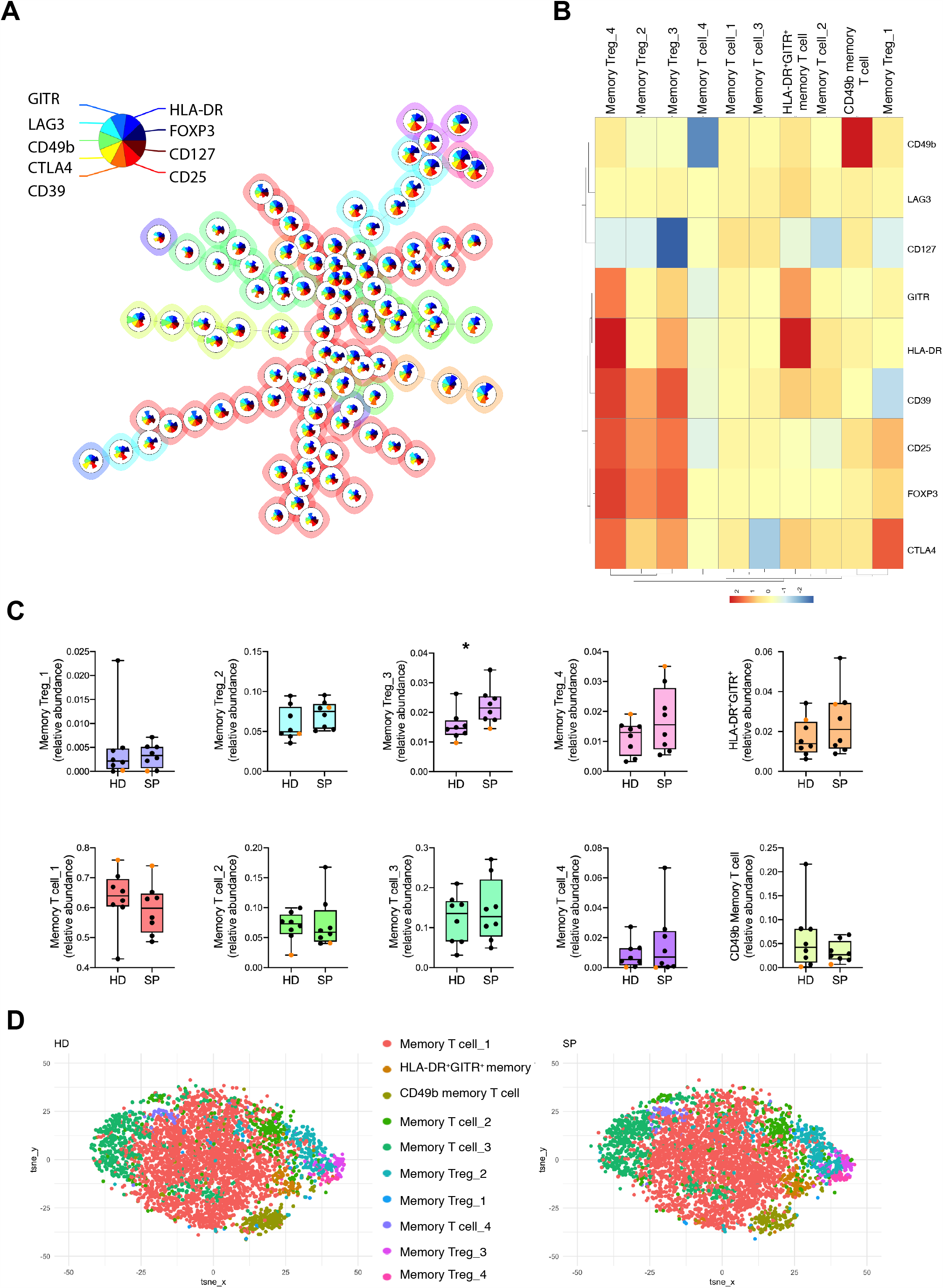
Expanded phenotypical analysis reveals that CD4 Tregs subtypes are increased in the SP cohort. Treg compartments generated by FlowSOM, clustering on live CD4^+^CD45RA^-^ cells from all donors. 10 metaclusters were identified based on marker expression: Memory T cell_1; Memory T cell_2; Memory T cell_3; Memory T cell_4; CD49b memory T cell; HLA-DR^+^GITR^+^ memory T cell; Memory Treg_1; Memory Treg_2; Memory Treg_3; Memory Treg_4. (A) Minimum Spanning Tree (MST) of 10 metaclusters generated using 9 different Treg markers. Each node represents a cluster (100 clusters) and larger metaclusters (10 metaclusters) are coloured around groups of nodes. Pie charts within each node represent expression levels of individual markers. (B) Heatmap with each metacluster to show overall marker expression. (C) Relative abundance boxplots for each metacluster identified for both healthy donor (HD) and slow progressor (SP) groups. (D) TSNE maps generated for HD and SP groups with overlays of each metacluster identified by FlowSOM. *<0.05, Wilcoxon matched-pairs signed rank test.

### Increased expression of GITR on an activated memory Treg population in slow progressors

Following the clustering analysis, we sought to determine if the memory T cell metaclusters, which we identified using FlowSOM, have a different phenotype in the slow progressor cohort, compared to healthy donors. Figure 3A shows expression heatmaps, with each marker used for clustering on each individual metacluster, for both SP and HD cohorts, not including donor 606. Unsurprisingly, the heatmaps revealed similar expression in each metacluster in both SP and HD; however, quantitative analysis demonstrated a significant increase in GITR expression in memory Treg_4 metacluster (Figure 3B, C), a metacluster with a high expression of GITR, HLA-DR, CD39, CD25, FoxP3, and CTLA4 (Figure 2, heatmap). Furthermore, we did not observe this GITR increase in donor 606, compared to the matched HD (Figure 3B, C, orange).

**Figure 3.**
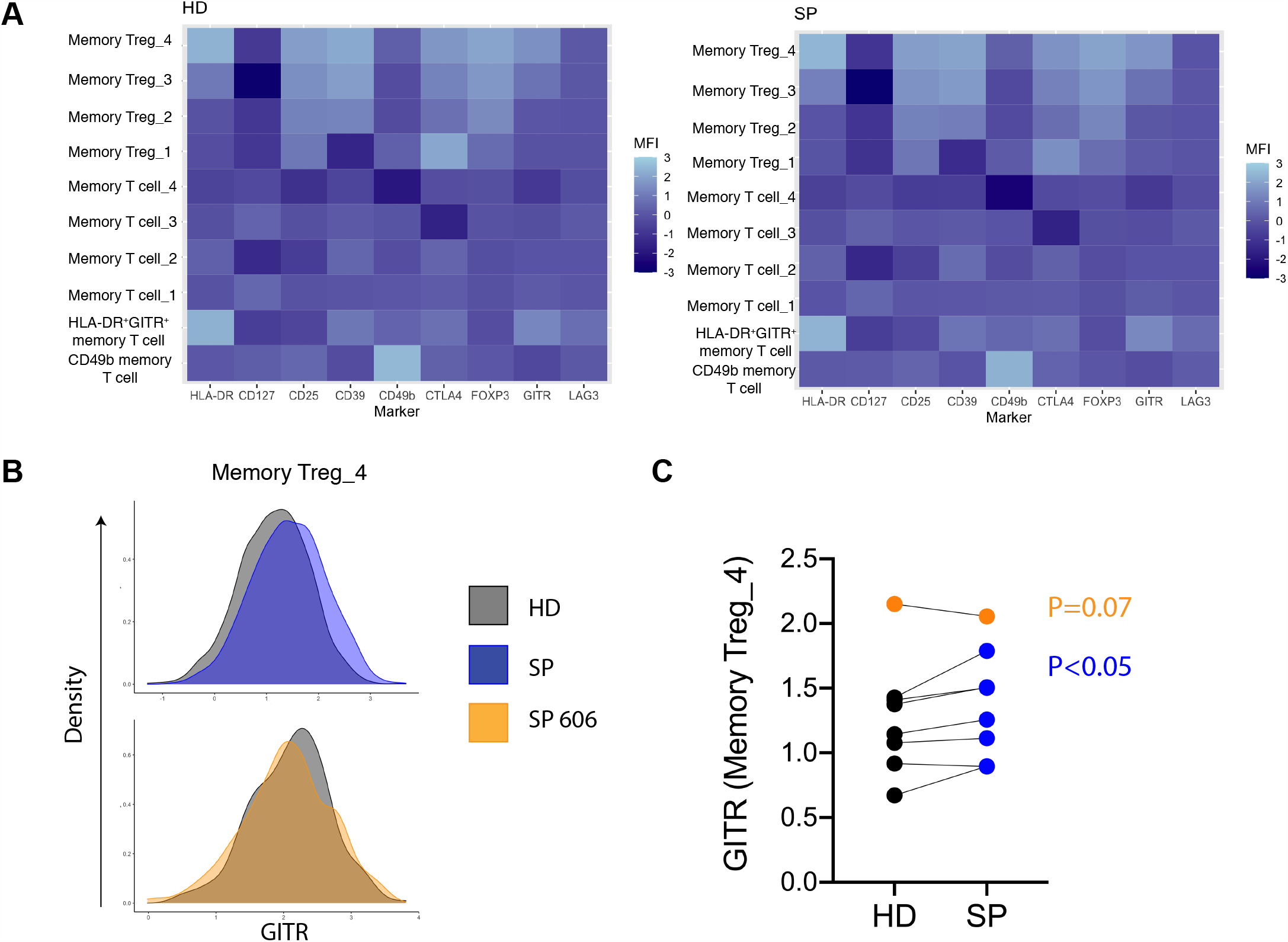
Increased GITR on memory Treg metacluster in slow progressor, compared to healthy donors. Each memory CD4 T cell metacluster (Memory T cell_1; Memory T cell_2; Memory T cell_3; Memory T cell_4; CD49b memory T cell; HLA-DR^+^GITR^+^ memory T cell; Memory Treg_1; Memory Treg_2; Memory Treg_3; Memory Treg_4), from FLOWSOM, was examined for a change in each expression marker. (A) expression heatmaps from both healthy donor (HD) and slow progressor (SP) cohort (SP 606 is not included). (B) Representative histograms showing GITR expression in HD (grey), SP (blue) and SP 606 (orange) in memory Treg_4 metacluster. (C) Summary graph for GITR expression in memory Treg_4. P<0.05, Wilcoxon matched-pairs signed rank test.

### Predictive modelling confirms increased Treg frequency is a signature of slow progressors

Following our FlowSOM analysis we utilised CITRUS (cluster identification, characterisation and regression), an algorithm that identifies different cellular signatures between grouped data (14) and provides predictive modelling. In this case, pre-gating was performed in Cytobank and clustered with the same cellular markers as our FlowSOM analysis. Analysis of CITRUS results provides a visualisation tree (Figure 4A). Here, SP and HD are clustered separately, based on marker expression - CD127, CD25 and FOXP3 are used to illustrate clustering of CD4 memory Tregs. Comparing our slow progressor cohort to the age- and gender-matched healthy controls, CITRUS identified 2 distinct clusters (false discovery rate 1%) that were different in frequency (Figure 4B). Analysis of cluster phenotypes (Figure 4C) revealed a similar phenotype to memory Treg 3 and memory Treg 4 metaclusters observed from FlowSOM analysis (Figure 2). Overall, high dimensional analysis confirms that increased frequency of CD4^+^ memory Tregs with a FOXP3^+^CD39^+^HLA-DR^+^GITR^+^CTLA4^+^ phenotype is a signature in slow progressors to type 1 diabetes.

**Figure 4.**
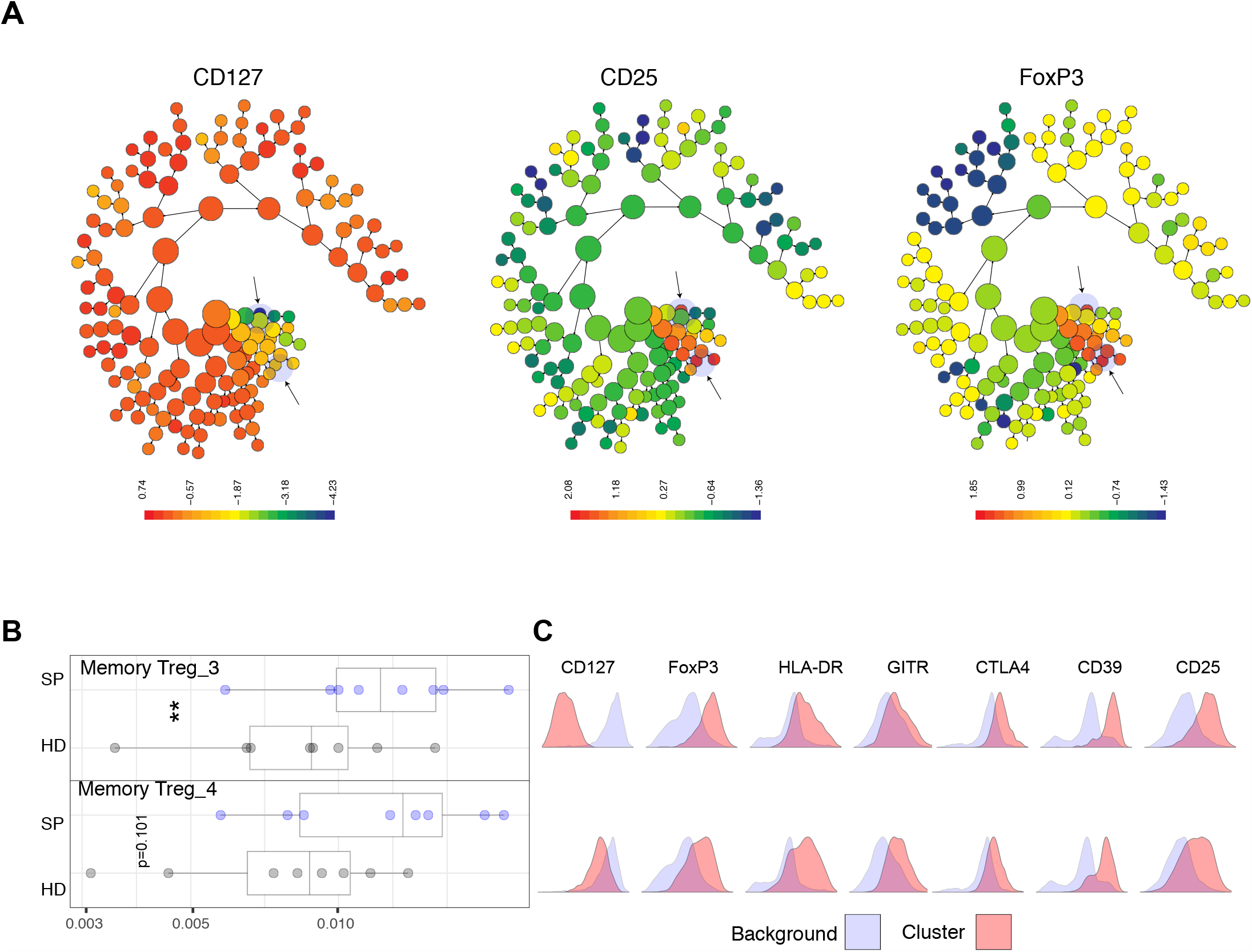
Predictive modelling using CITRUS confirms increased Treg frequency is a signature of slow progressors. CITRUS analysis was performed comparing slow progressors and matched healthy donors on CD4^+^CD45RA^-^ T cells. (A) CITRUS coloured by channels CD127, CD25 and FOXP3, clusters identified as different in slow progressor cohort highlighted by arrows. (B) Boxplots to show relative abundance of CITRUS memory Treg_3 and memory Treg_4 in slow progressors (SP, blue dots) and healthy donors (HD, black dots). (C) Histograms to show phenotype of each cluster (pink) and relative expression of Treg markers compared to background expression (blue). **<0.01, Wilcoxon matched-pairs signed rank test.

## Conclusions

Our findings demonstrate that individuals who have been positive for multiple islet autoantibodies for 10 years or more, and have not developed type 1 diabetes, have a unique Treg signature. This signature is characterised by impaired Tregs, in terms of effector CD4^+^ T cell suppression; however, effector CD4^+^ T cells in the slow progressors are more responsive to suppression. Furthermore, we find activated memory Tregs are increased in frequency in slow progressor individuals, compared to age and sex-matched healthy donors. These activated memory Tregs are enriched in GITR expression. Our study also features a unique snapshot of a slowly progressing individual (606), assessed at a time of raised HbA1c, diagnostic of development of type 1 diabetes, and this individual’s response was altered compared to the slow progressor cohort. Interestingly, no loss of Treg function was observed in this individual, but the effector CD4^+^ T cells were resistant to suppression, specifically CD4^+^ T cell activation, which was accompanied by an increase in IFNγ and IL-17 in Treg co-cultures, compared to the control. In terms of Treg numbers, this former slow progressor, maintained the increase in Treg frequency, but a similar expression of GITR, compared to the matched control.

Functional Treg studies in type 1 diabetes cohorts has demonstrated both impaired suppressive function in Tregs and a resistance to suppression in effector T cells. Together, both are likely to contribute to the impaired suppressive action, which varied in different individuals, and so a certain level of heterogeneity may exist in individuals with type 1 diabetes (5; 15). Here, we demonstrate both impaired Treg function in parallel with an increased responsiveness to suppression in slow progressors. However, it is interesting that despite an impairment of Tregs in slow progressors, these individuals remain free of diabetes, yet in the donor, on the threshold development of diabetes, this impairment was not observed. Equally, we find the opposite pattern with effector T cells, with an increased response to suppression in slow progressors, which was lost in donor 606. The resistance to suppression in donor 606 was associated with an increase in pro-inflammatory cytokines, and therefore it is possible that Tregs, were responding to pro-inflammatory cytokines with increased activity under certain circumstances, IFNγ is required for Treg mediated suppression (16). Furthermore, the Tregs may also contribute to the increased IFNγ and IL-17A detected, as Tregs that are Th1 and Th17-like have been identified (17), and these Treg produced pro-inflammatory cytokines, may be required for suppressive activity (17; 18). Our study highlights the crucial interplay between Treg and effector T cells, and it will be important to understand the role and timing of Treg dysfunction during disease progression.

Unsupervised clustering allowed us to profile memory Tregs in slow progressors, which revealed multiple Treg subsets with varying degrees of expression of markers associated with antigen experience and mechanisms of suppression. We observed an increase in activated memory Tregs in slow progressors, compared to matched controls. Overall, Tregs in type 1 diabetes cohorts are similar in frequency to control individuals (15), although a recent study reported a reduction in activated FoxP3 Tregs in type 1 diabetes (19). Currently, we cannot determine whether the subsets we have identified represent Tregs at differing points in maturity or differentiation, or whether they are distinct subsets. Additional Treg markers may be required for the identification of other Treg subsets (12) in this distinctive group of slow progressor individuals and this is for further study.

Interestingly, we observed an increase in the expression of GITR, a member of the TNFR family, in a distinct Treg metacluster, in slow progressors. GITR expression, which is frequently observed on activated Tregs (20) and is critical for suppressive action in Tregs, shown by the use of anti-GITR antibodies (20; 21). However, in some settings using anti-GITR agonistic antibodies has not affected the suppressive ability of Tregs, but rather induced proliferation and migration of pathogenic T cells (22). Indeed, reduced expression of GITR on Tregs or a loss in GITR^+^ Tregs in individuals with type 1 individuals does not result in an impaired suppressive phenotype, but the Tregs are more susceptible to apoptosis, compared to healthy donors (23). In mouse models, GITR is crucial for the expansion of Tregs, as demonstrated both in GITR-knockout mice, which have a reduced Treg frequency (24), as well as in GITR-L deficient mice that demonstrate impaired Treg expansion (25). These studies suggest that increased expression of GITR on Tregs in slow progressors may be the result of an increased propensity to develop or expand. Further functional studies are required to identify if activated memory GITR^+^ Tregs in slow progressors have an improved survival mechanism or a resistance to apoptosis. Considering the importance of the GITR/GITR-L pathway, in both effector T cells and Tregs, it would also be important to identify any difference in GITR-L signatures in immune cell compartments known to express this ligand.

A limitation in this study was a lack of age-matched individuals with new-onset type 1 diabetes. It is a challenge to match our slow progressors to people with new-onset diabetes, as they were considerably older than many newly diagnosed individuals with type 1 diabetes having had autoantibodies, but not developed diabetes, for many years. For the future, it would also be beneficial to profile the Treg responses and signatures in ‘at-risk’ individuals positive for multiple islet autoantibodies, allowing us to track and predict the individuals who will slowly progress, in real-time. Overall, our study describes a rare case of the islet autoantibody and Treg characteristics at diagnosis of type 1 diabetes in an older adult. Furthermore, we identify an immune cell signature in extreme slow progressors and our study highlights the need to develop our understanding of Treg heterogeneity that exists during the development of type 1 diabetes, in order to help stratify those who would benefit from regulatory T cell therapy.

## Data Availability

Data available on request.

## Acknowledgements

The BOX study was funded by Diabetes UK (14/0004869). The SNAIL study was funded by a Strategic Research Agreement from the JDRF (17-2013-529). AEL was funded by a Diabetes UK/JDRF RD Lawrence Fellowship (18/0005778 and 3-APF-2018-591-A-N). KG and FSW are the guarantors of this work and, as such, had full access to all the data in the study and take responsibility for the integrity of the data and the accuracy of the data analysis. No conflict of interest exists for this study.

## Author contributions

JB, AL, KG and FSW designed the experiments and wrote the manuscript. JB performed the experiments and analysed the data. SJH contributed to data analysis. RJA, IVW and CM supported data acquisition. All authors reviewed the manuscript.

