## Supplementary data for "Unsupervised clustering reveals a unique Treg profile in slow progressors to type 1 diabetes"

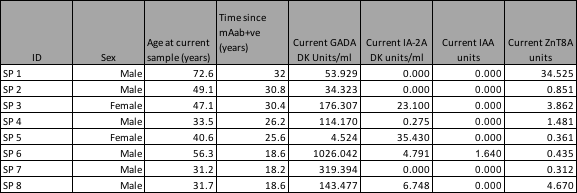

**Table 1.** Current islet autoantibody results for 8 slow progressors. The positive thresholds for GADA are >=33 DK units/ml, IA-2A are >=1.4 DK units/ml, IAA are >=0.2 units and ZnT8A are >=1.8 units.

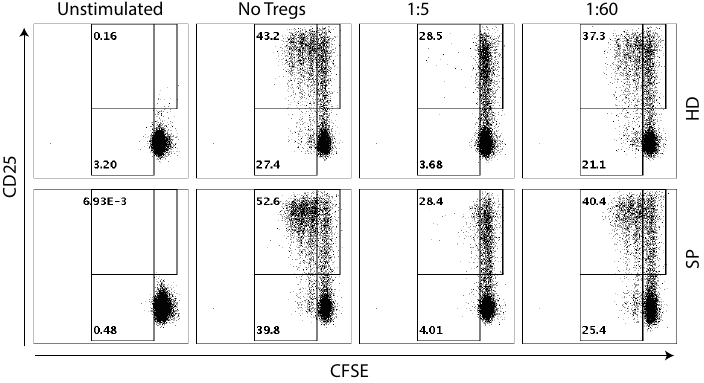

*Supplementary Figure 1. Representative flow plots for Treg suppression assay.* CD4 Treg suppression assay flow plots showing CFSE and CD25 gating on both slow progressor donor (SP) and matched healthy donor (HD). Unstimulated control, no Tregs (positive control), Treg: responder ratios 1:5 and 1:60 is shown.

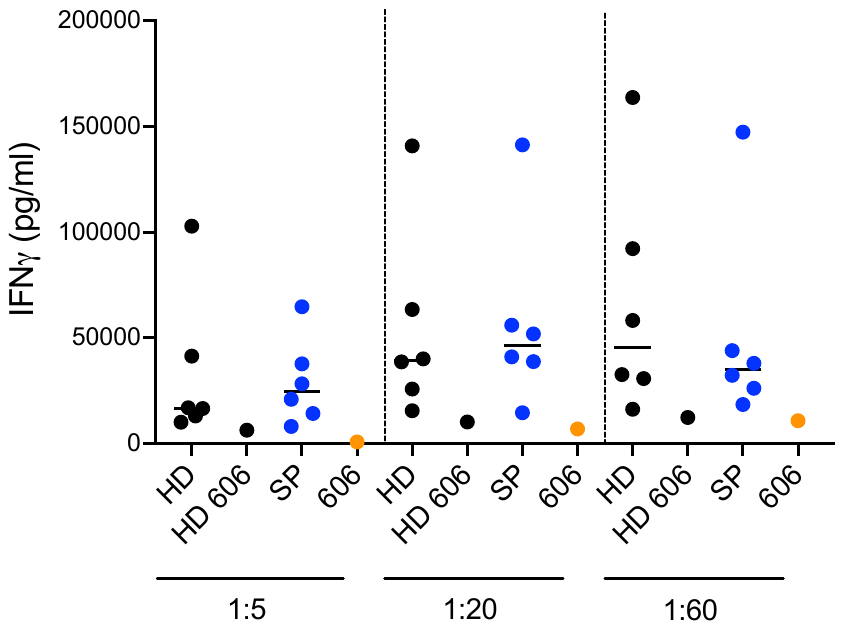

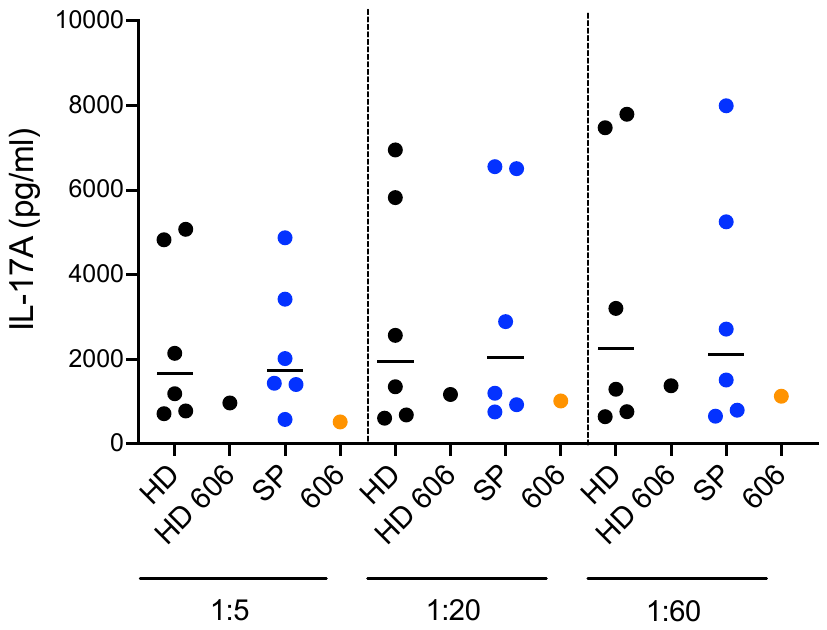

*Supplementary Figure 2. Pro-inflammatory cytokine expression from Treg suppression assays.* HD responders were co-cultured with Tregs from matched healthy donors (HD, black dots, HD 606 plotted separately) and slow progressors (SP, blue dots) or SP 606 donor (orange dots). IFNγ (left) and IL-17A (right) production at different Treg:responder ratios, stated below the graph measured by Meso Scale Discovery.

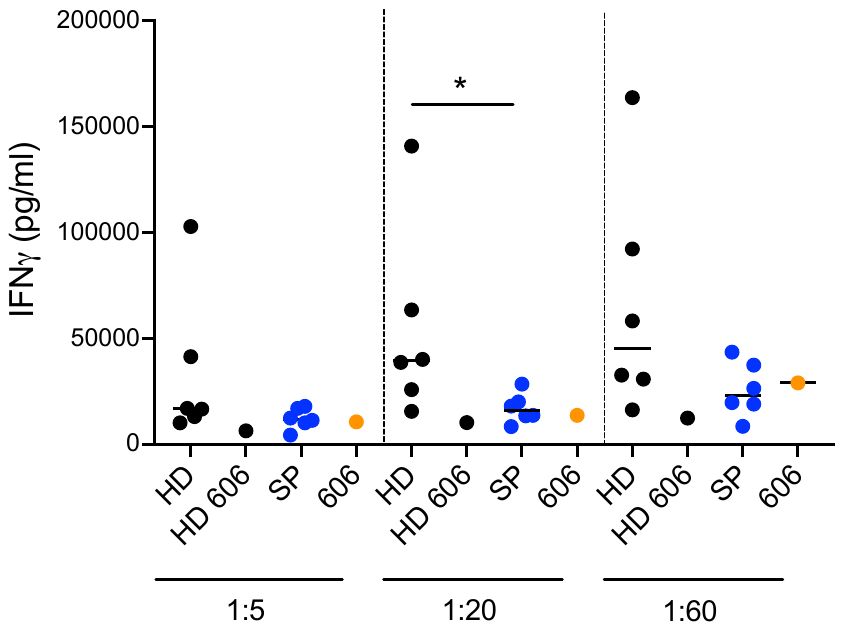

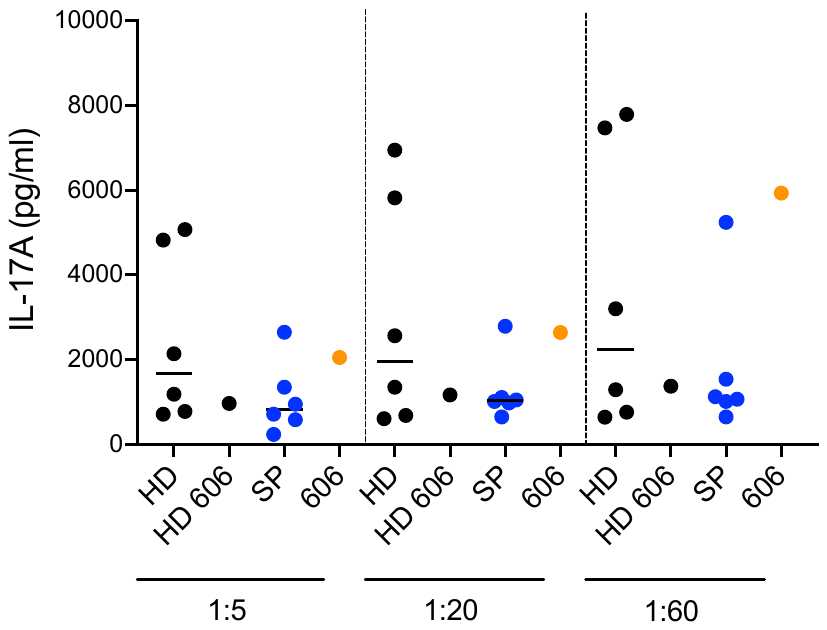

*Supplementary Figure 3. Pro-inflammatory cytokine expression from autologous Treg suppression co-cultures.* IFNγ (left) and IL-17A (right) production at different Treg: responder ratios, stated below the graph, in healthy donors (black dots, HD 606 plotted separately), slow progressors (blue dots) and SP 606 donor (orange dots) measured by Meso Scale Discovery.

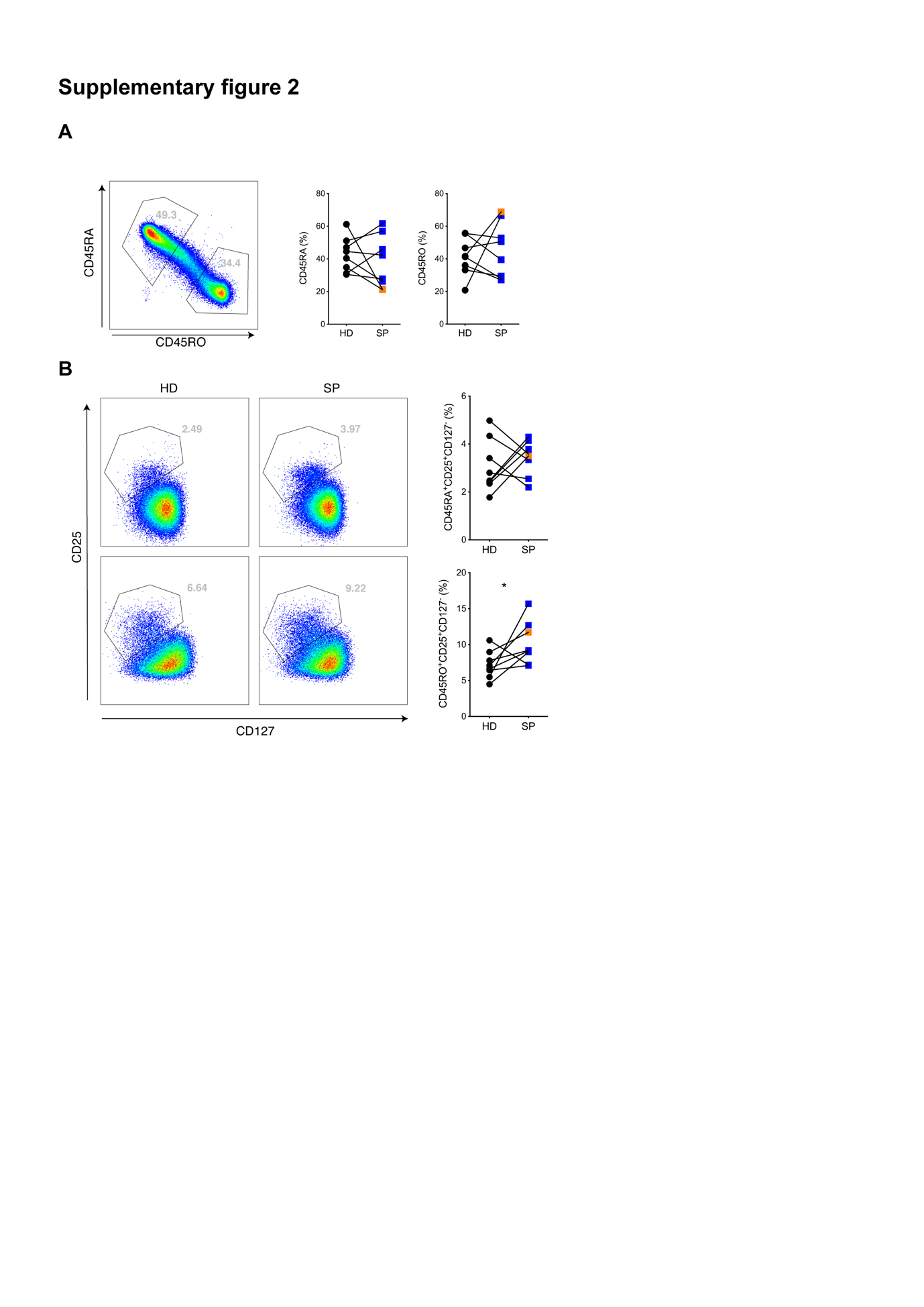

*Supplementary Figure 4. Manual gating of resting naïve and memory Tregs* (A) flow cytometric plots and summary graphs showing expression of CD4^+^CD45RA^+^ and CD4^+^CD45RO^+^ T cells in healthy donors (black dots) and slow progressors (blue squares) and donor 606 (orange square). (B) flow cytometric plots and summary graphs to show CD4+CD25+CD127- Tregs in both CD45RA (naïve) and CD45RO (memory) subsets in healthy donors (black dots) and slow progressors (blue squares) and donor 606 (orange square). *<0.05, Wilcoxon matched-pairs signed rank test.
